# Deficient goal-directed control in a population characterized by extreme goal pursuit

**DOI:** 10.1101/19002089

**Authors:** Karin Foerde, Nathaniel D. Daw, Teresa Rufin, B Timothy Walsh, Daphna Shohamy, Joanna E. Steinglass

## Abstract

Computational neuroscience has contributed to understanding compulsive behavior by distinguishing habitual from goal-directed choice through model-free and model-based learning. Yet, questions remain about applying this approach to psychiatric conditions that are characterized by complex behaviors that occur outside the laboratory. Here, we compared individuals with anorexia nervosa (AN), whose self-starvation appears both excessively goal-directed *and* habitual, with healthy controls (HC) to assess: 1) whether their behavior is characterized by enhanced or diminished model-based behavior, 2) the domain specificity of any abnormalities by comparing learning in a food-specific context as well as in a monetary context, and 3) whether impairments are secondary to starvation by comparing learning before and after initial treatment. Across all conditions, individuals with AN showed an impairment in model-based, but not model-free, learning, suggesting a general and persistent contribution of habitual over goal-directed control, across domains and timepoints.

## Introduction

The pathophysiology of psychiatric illnesses is poorly understood. Little is known about etiologies, diagnostic tests mainly serve to rule out medical illness, and underlying neural mechanisms remain mysterious. These problems stem, in part, from the great challenge of understanding the mechanisms of mental processes even in the healthy brain. Recent advances in cognitive and computational neuroscience offer promising new approaches for understanding brain mechanisms underlying psychiatric symptoms (Huys, Maia, & Frank, 2016). For example, the emerging understanding of the brain’s systems for habitual vs. goal-directed control has offered a potential mechanism underlying compulsive symptomatology (Everitt & Robbins, 2016), which can occur across a range of psychiatric illnesses. In the present study, we apply methods for probing habitual versus goal-directed behavior in relation to anorexia nervosa (AN), in which the central behavioral feature appears to be both extremely goal-focused (unrelenting pursuit of thinness) and yet rigidly unchangeable (poor responses to treatment) (Walsh, 2013), thereby challenging a simple automatic-vs-controlled dichotomy.

The existence of a dichotomy between automatic and controlled behavior is a long-standing organizing principle in psychology and neuroscience (James, 1890), and recent research shows that controlled or goal-driven, motivated behaviors can be distinguished behaviorally and neurally from cue-elicited, automatic behaviors (“habits”) (Dickinson & Balleine, 2002; Graybiel, 2008). These two types of behavior have been argued to arise from distinct computational mechanisms for evaluating actions, known as model-based and model-free learning (Daw, Gershman, Seymour, Dayan, & Dolan, 2011; Daw, Niv, & Dayan, 2005). Goal-directed (or model-based) behavior enables pursuit of long-term goals, even if there is conflict with short-term temptations, and its deliberative nature offers the ability to update behavior when the value of an outcome changes. Habitual (or model-free) behavior results from repetition of stimulus-response associations and facilitates efficient automatic behavior in response to cues but renders behavior relatively inflexible. In the lab, these approaches can be distinguished using instrumental learning tasks that include outcome devaluation procedures and two-step Markov decision tasks, and are associated with partially distinct neural substrates (Coutureau & Killcross, 2003; Daw et al., 2011; Glascher, Daw, Dayan, & O’Doherty, 2010; Killcross & Coutureau, 2003; Lee, Shimojo, & O’Doherty, 2014; Tricomi, Balleine, & O’Doherty, 2009; Valentin, Dickinson, & O’Doherty, 2007; Yin, Knowlton, & Balleine, 2004; Yin, Knowlton, & Balleine, 2006).

Experimental evidence shows that healthy people generally use a mix of both model-free and model-based approaches, shifting the balance between them depending on circumstances (Daw et al., 2011; Otto, Gershman, Markman, & Daw, 2013; Otto, Raio, Chiang, Phelps, & Daw, 2013; Weissengruber, Lee, O’Doherty, & Ruff, 2019). In contrast, reductions in model-based, goal-directed behavior have been shown in disorders with compulsive behaviors, such as obsessive compulsive disorder (OCD) and binge eating disorder (Gillan et al., 2015; Gillan, Robbins, Sahakian, van den Heuvel, & van Wingen, 2016; Voon et al., 2015). Together, these studies suggest that a computational approach that distinguishes the contributions of habitual and goal-directed behavior may help link psychiatric symptoms with dysfunction in basic neural mechanisms.

Compulsive behavior, broadly, is action engaged in persistently, even when unrewarding or maladaptive, and often despite an individual’s own assessment that it is excessive. Multiple psychiatric disorders are characterized by behaviors with a seemingly compulsive character (e.g., tic disorders, obsessive-compulsive disorder, substance use disorders, and eating disorders; American Psychiatric Association, 2013) and the persistence of these varied compulsive behaviors is puzzling. One hypothesis is that excessive relative dominance of automatic behavior underlies compulsivity (Everitt & Robbins, 2016; Gillan et al., 2015; Gillan & Robbins, 2014). However, some basic questions remain about the adequacy and explanatory potential of this framework for elucidating mechanisms of psychiatric illness, three of which we address in the current study.

First, a range of psychiatric illnesses include behaviors that can be considered compulsive, but the behaviors themselves are not always easily captured by the simple, repetitive, stimulus-evoked habits described in the experimental literature. That is, categorizing the real-world behaviors at the core of psychiatric illnesses is complex. For example, engagement in a ritualized behavior to reduce anxiety symptoms in OCD may, in some ways, be compulsive and automated, yet it can also be construed as a successful, goal-directed, strategy for managing anxiety due to obsessions (Salkovskis, 1985). Similarly, drug-seeking behavior may be compulsive, and at the same time require considerable flexible goal pursuit (Tiffany, 1990).

AN provides a particularly intriguing example. Maintenance of significantly low body weight is a defining feature of AN (American Psychiatric Association, 2013). This self-starvation appears to be an unrelenting goal pursuit focused on weight loss, and individuals with AN are commonly thought of as rigidly self-controlled (Bruch, 1979). Data from temporal discounting studies show that individuals with AN are more likely than healthy comparison participants to forego immediate monetary rewards in favor of larger, delayed rewards during intertemporal choices, consistent with descriptions of heightened self-control (Decker, Figner, & Steinglass, 2015; Steinglass et al., 2012; Steinglass et al., 2017). And yet, the restrictive eating that characterizes AN also shares many features of habit (Foerde, Steinglass, Shohamy, & Walsh, 2015; Walsh, 2013): it is learned (not innate), is inflexibly triggered by certain cues, and individuals with AN are unable to readily change this behavior—even when seeking treatment, suggesting that it has become outcome-independent over time. The possibility that persistent maladaptive choices in AN may reflect overlearned habits is particularly intriguing given that the behavior is characterized by choices against normatively rewarding foods (e.g. foods high in sugar and fat). Thus, it remains unclear whether maladaptive behavior in AN reflects excessively goal-directed or excessively habitual behavioral control. More operationally, the question remains: if behavior in AN arises from imbalance between goal-directed and habitual mechanisms, which one is dominant?

Second, goal-directed and habitual behavior are domain-general mechanisms, as illustrated by their trans-diagnostic application, yet, different psychiatric illnesses involve quite specific compulsive behaviors (e.g., OCD with specific compulsions, abuse of specific substances, gambling, or excessive weight loss). Most, if not all, laboratory studies have used generic outcomes, typically money (Gillan et al., 2016; Voon et al., 2015) or food (Tricomi et al., 2009; Valentin et al., 2007), to study goal-directed and habitual behavior. An important interpretational question left open by these studies is whether apparent deficits in goal-directed control in psychiatric patients are due to impairments in a domain-general mechanism, or instead to a reorienting of goal-directed behavior toward the object of compulsion at the expense of other domains. The latter view would predict that substance abusers, for instance, are relatively unmotivated by money (and unwilling to pursue it with goal-directed control) but could mitigate or even reverse their impairment if their drug of abuse was at stake. Similarly, for individuals with AN, the balance of model-based and model-free control may be different for choices about money than for choices about food. Thus, does psychopathology reflect a domain-general deficit in goal-directed (model-based) learning, or something more outcome-specific? Whereas studying many illness-specific outcomes (including drugs of abuse) in the lab comes with obvious challenges, studying AN offers an opportunity to address this question via tasks in the setting of food outcomes compared with money outcomes.

Third, the presence of model-based deficits in the setting of psychiatric illness only indicates a correlation between the two and does not directly support the presumption that imbalanced learning is a causal factor in developing maladaptive compulsions. Indeed, decreased goal-directed control may be a *result* of compulsive behaviors, as has been argued, for instance, for neurological effects of drugs of abuse (Volkow et al., 2010). Overall, little is known about the longitudinal progression of these deficits. For instance, are deficits in goal-directed control remediated by treatment of psychopathology in disorders of compulsion? If deficits are remediated, this would suggest that they are an acute symptom of the disorder, and likely not a pre-existing cause. A recent study suggested that impairments in goal-directed behavior in OCD are not remediated by cognitive behavioral therapy (Wheaton, Gillan, & Simpson, 2018). For AN particularly, the secondary effects of starvation on cognition might be substantial. Thus, by studying individuals with AN during acute illness and again after extensive inpatient weight-restoration treatment, the persistence of any learning differences and their relation to changes in psychopathology can be assessed.

In this study we addressed these questions with a population of individuals with AN using a two-step Markov decision task (**Figure 1**) designed to assess the extent of model-based vs. model-free learning. By examining an illness with complex and seemingly highly controlled (yet maladaptive) behaviors, we aimed to test whether these reflect heightened or deficient model-based behavior. We also compared results from food and monetary versions of the task to examine the specificity of findings in the eating disorder domain and tested for the presence of model-based vs. model-free behavior in the context of outcomes directly relevant to underlying psychopathology. Whereas the monetary task assumes that money is rewarding, in the food task participants were rewarded with points that they could use to select the food of their choice, thereby allowing the food-relevant task to be motivationally relevant to both controls and patients. Finally, we evaluated patients before and after inpatient weight-restoration treatment, to begin to study the progression of model-based and model-free processes.

**Figure 1.**
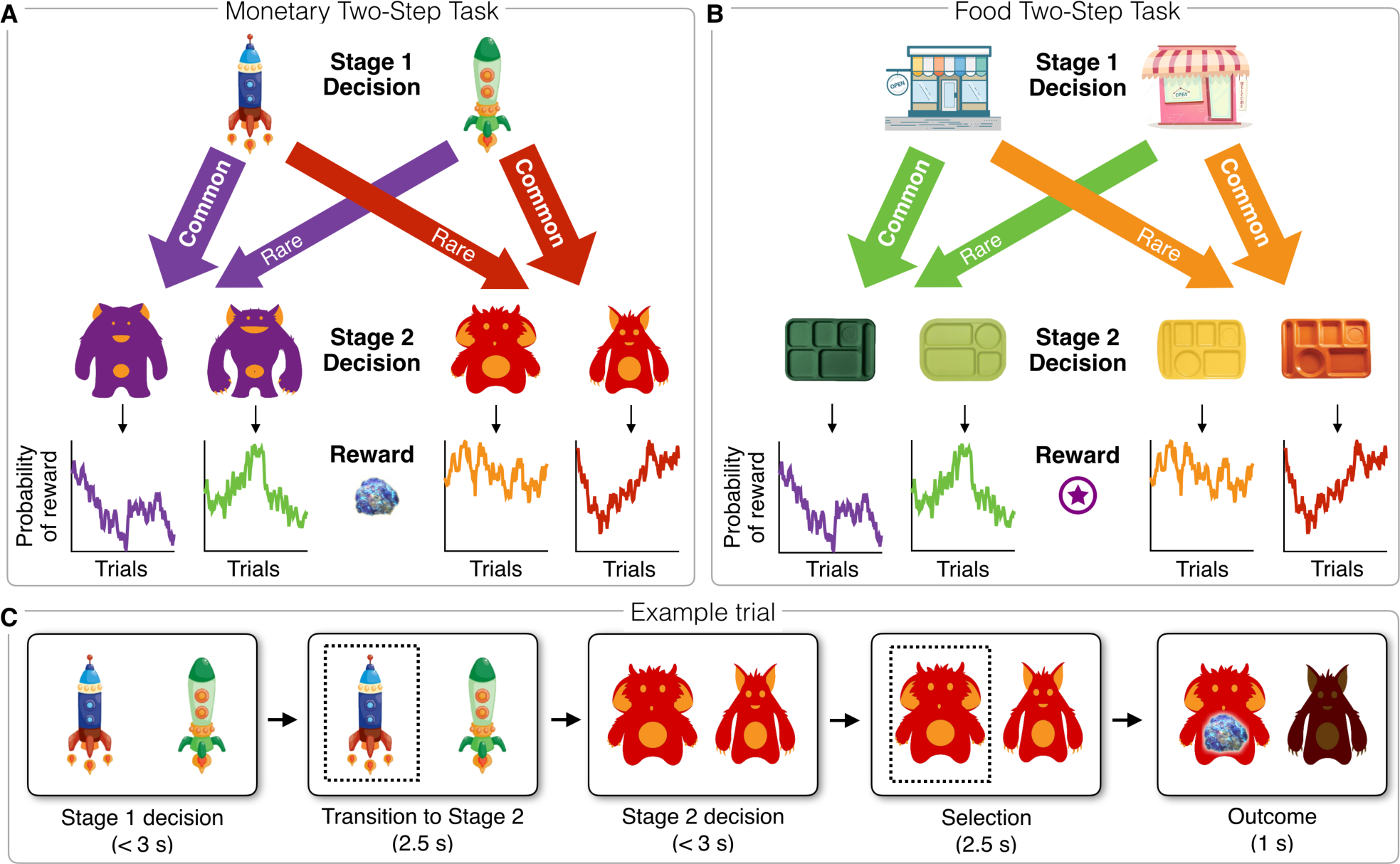
Two-step decision tasks used to assess model-free and model-based learning. **A**) Decision task with monetary outcomes. Alien treasure pieces were converted into a monetary bonus at the end of the task. **B**) Task with food outcomes. Food tokens were converted into access to a range of preferred snack items and the selected snack was consumed after the task. (**A, B**) In both tasks, the Stage 1 choice determined the transition to the next stage according to a fixed probability. One choice was associated with transition to one particular Stage 2 state 70% of the time (Common transition) and the other state 30% of the time (Rare transition). At Stage 2, participants made choices followed by reward or no reward (both monetary and food outcomes were actualized after the task). Each Stage 2 option was associated with a probabilistic reward, which ranged from 0.25-0.75 and varied gradually (according to a Gaussian random walk) and independently across trials (see examples in bottom rows of **A** and **B**). **C**) Example trial steps from the monetary task.

## Results

As expected, the HC and AN groups differed on measures of eating disorder severity (Eating Disorder Examination Questionnaire, EDEQ; (Fairburn, 2008; Fairburn & Beglin, 1994)), depression (Beck Depression Inventory, BDI; (Beck & Steer, 1993)), anxiety (State-Trait Anxiety Inventory, STAI; (Spielberger, Gorsuch, & Lushene, 1970)), and body mass index (BMI) at Time 1, but did not differ on age (see **Table 1**). Although all participants had IQ scores in the normal range, there was a statistically significant group difference; IQ (which has been associated with model-based learning in previous studies; (Gillan et al., 2016)) was included as a covariate in all analyses. Additionally, age was included as a covariate, as the sample spanned adolescents and adults and use of model-based learning has been shown to develop gradually across adolescence (Decker, Otto, Daw, & Hartley, 2016). Mean BMI increased significantly from Time 1 to Time 2 among individuals with AN (t(23)=-13.01, p < 0.001) and did not differ significantly between groups at Time 2, indicating successful weight restoration among individuals with AN. In addition, among individuals with AN, the expected psychological change was seen as measures of eating disorder pathology (EDEQ: t(24)=6.7, p < 0.001), depression (BDI: t(23)=5.15, p < 0.001), and anxiety (STAI: t(24)=3.36, p = 0.003), significantly improved from Time 1 to Time 2, although these measures remained elevated relative to HC (ps < 0.001, see **Table 1**).

**Table 1.** Demographic and clinical information

|  | Time 1 |  |  |  | Time 2 |  |  |  |
| --- | --- | --- | --- | --- | --- | --- | --- | --- |
|  | HC (n=53) | AN (n=41) | *t* | *p* | HC (n=29) | AN (n=25) | *t* | *p* |
| | Mean ± SD | Mean ± SD | | | Mean ± SD | Mean ± SD | | |
| Age (years) | 25.6 ± 5.0 | 27.1 ± 7.0 | 1.2 | 0.236 | | | | |
| WASI Est. IQ | 111.8 ± 12.2 | 105.1 ± 11.3 | -2.7 | 0.008 | | | | |
| BMI (kg/m<sup>2</sup>) | 21.3 ± 1.5 | 16.0 ± 2.0 | -14.6 | < 0.001 | 20.9 ± 1.3 | 20.4 ± 0.9 | -1.4 | 0.172 |
| Dur. Ill. (years) | n/a | 9.6 ± 7.1 | | | | | | |
| EDEQ | 0.5 ± 0.6 | 4.16 ± 1.5 | 15.8 | < 0.001 | 0.4 ± 0.4 | 2.8 ± 1.2 | 9.8 | < 0.001 |
| BDI | 2.1 ± 2.4 | 27.6 ± 12.6 | 14.4 | < 0.001 | 3.0 ± 2.6 | 18.0 ± 14.0 | 5.7 | < 0.001 |
| STAI (T) | 31.3 ± 7.3 | 61.0 ± 11.2 | 15.5 | < 0.001 | 33.6 ± 8.7 | 56.4 ± 12.4 | 7.9 | 0.003 |
| YBC-EDS Total | n/a | 22.5 ± 6.4 | | | | | | |
| Preoccupation | n/a | 11.7 ± 3.2 | | | | | | |
| Rituals | n/a | 10.8 ± 3.7 | | | | | | |

AN: anorexia nervosa; BDI: Beck Depression Inventory; BMI: body mass index; Dur. Ill: duration of illness; EDEQ: Eating Disorder Examination questionnaire; HC: healthy comparison participant; STAI(T): State-Trait Anxiety Inventory; YBC-EDS: Yale-Brown-Cornell Eating Disorder Scale; WASI: Wechsler Abbreviated Scale of Intelligence. WASI IQ data are missing from 1 AN. At Time 1, BDI data are missing from 1 AN. At Time 2, BMI data are missing from 1 AN.

To examine task performance, we first tested whether individuals with AN differed from HC in model-free and model-based learning overall, across both monetary and food outcome versions at both assessment points (modeling any effects of task and session but considering first the main effect of group). There was a significant contribution of model-free learning to behavior in both groups (ps < 1e-5) and no significant difference between groups (Est: 0.09, SE = 0.06, z = 1.36, p = 0.174; **Fig. 2**). In contrast, model-based learning was also present in both groups (ps < 1e-5) but significantly attenuated in the AN group relative to the HC group (Est: 0.15, SE = 0.06, z = 2.27, p = 0.023; **Fig. 2**). To test the specificity of the group difference to model-based relative to model-free learning, we repeated the analysis using the parameter *w* (see Methods), which measures the relative reliance on model-based learning and found that the relative reliance *w* was increased among HC (Est: 0.41, SE = 0.2, z = 2.06, p = 0.039).

**Figure 2:**
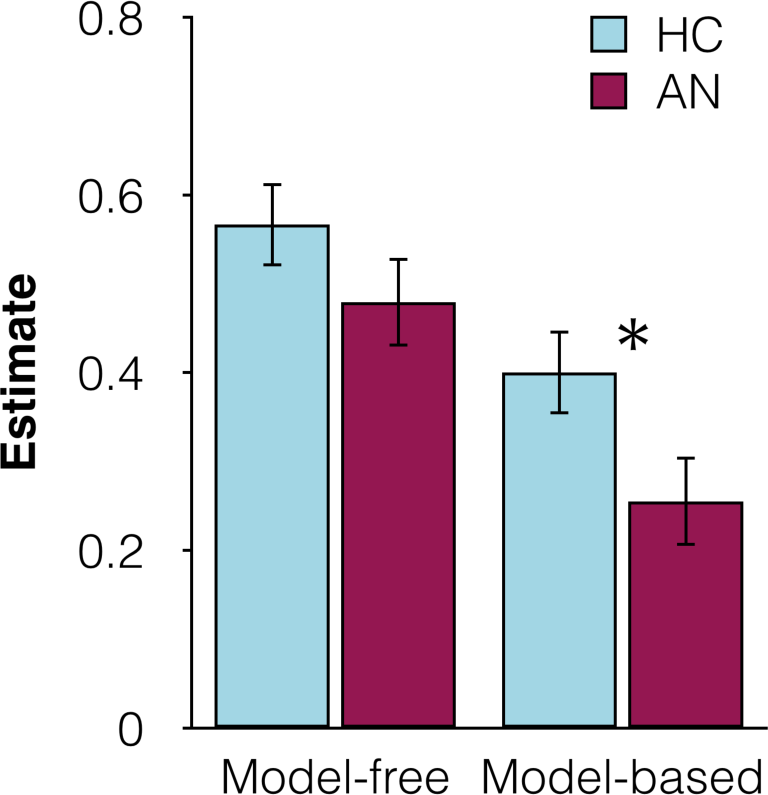
Overall model-free and model-based contributions to learning for HC and AN across task type (monetary and food) and session (Time 1 and Time 2).

Next, we tested whether the model-based deficit was domain-general or specific to monetary vs. food outcomes (**Table 2**). Task type did not interact significantly with group differences in model-based learning (Est: -0.08, SE = 0.07, z = -1.11, p = 0.27) and the same pattern of impaired model-based learning in AN was observed regardless of outcome (**Fig. 3A**). There was, however, a main effect of task such that all participants showed a greater model-based contribution in the monetary task relative to the food outcome task (Est: 0.15, SE = 0.05, z = 2.78, p = 0.006).

**Table 2.**
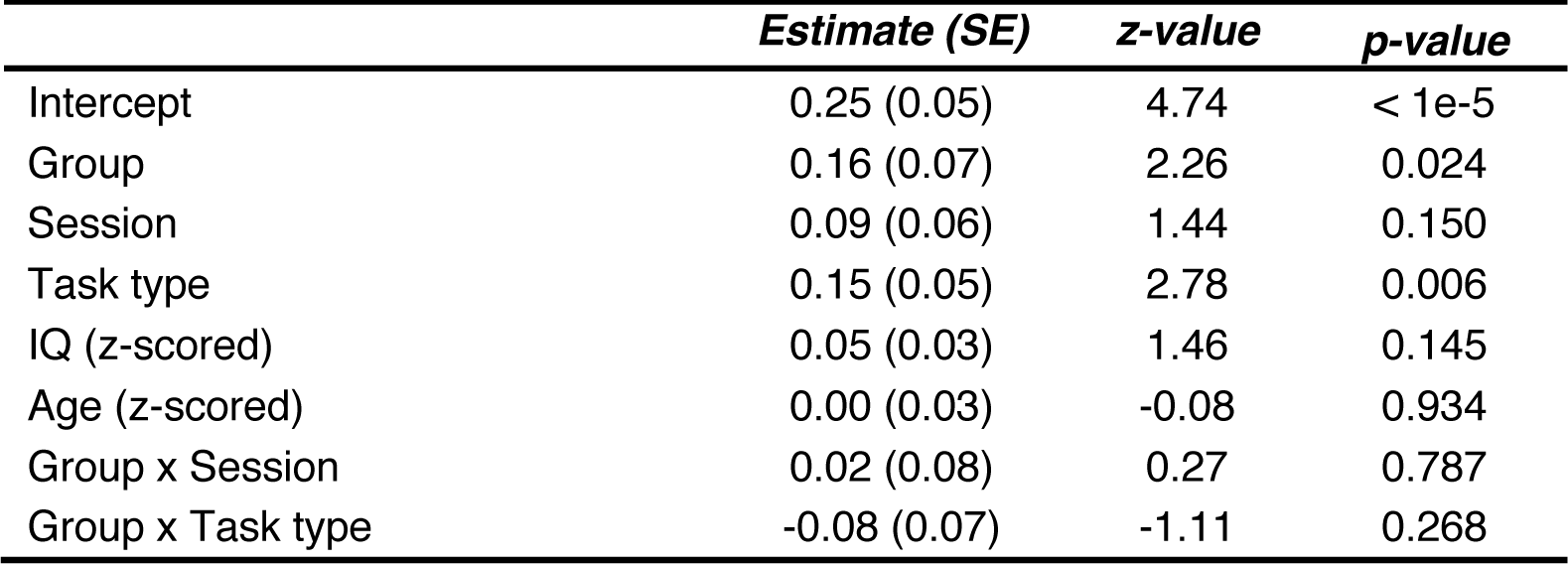
Model based learning: regression analysis including Group, Session, and Task type

|  | *Estimate (SE)* | *z-value* | *p-value* |
| --- | --- | --- | --- |
| Intercept | 0.25 (0.05) | 4.74 | < 1e-5 |
| Group | 0.16 (0.07) | 2.26 | 0.024 |
| Session | 0.09 (0.06) | 1.44 | 0.150 |
| Task type | 0.15 (0.05) | 2.78 | 0.006 |
| IQ (z-scored) | 0.05 (0.03) | 1.46 | 0.145 |
| Age (z-scored) | 0.00 (0.03) | -0.08 | 0.934 |
| Group x Session | 0.02 (0.08) | 0.27 | 0.787 |
| Group x Task type | -0.08 (0.07) | -1.11 | 0.268 |

**Figure 3:**
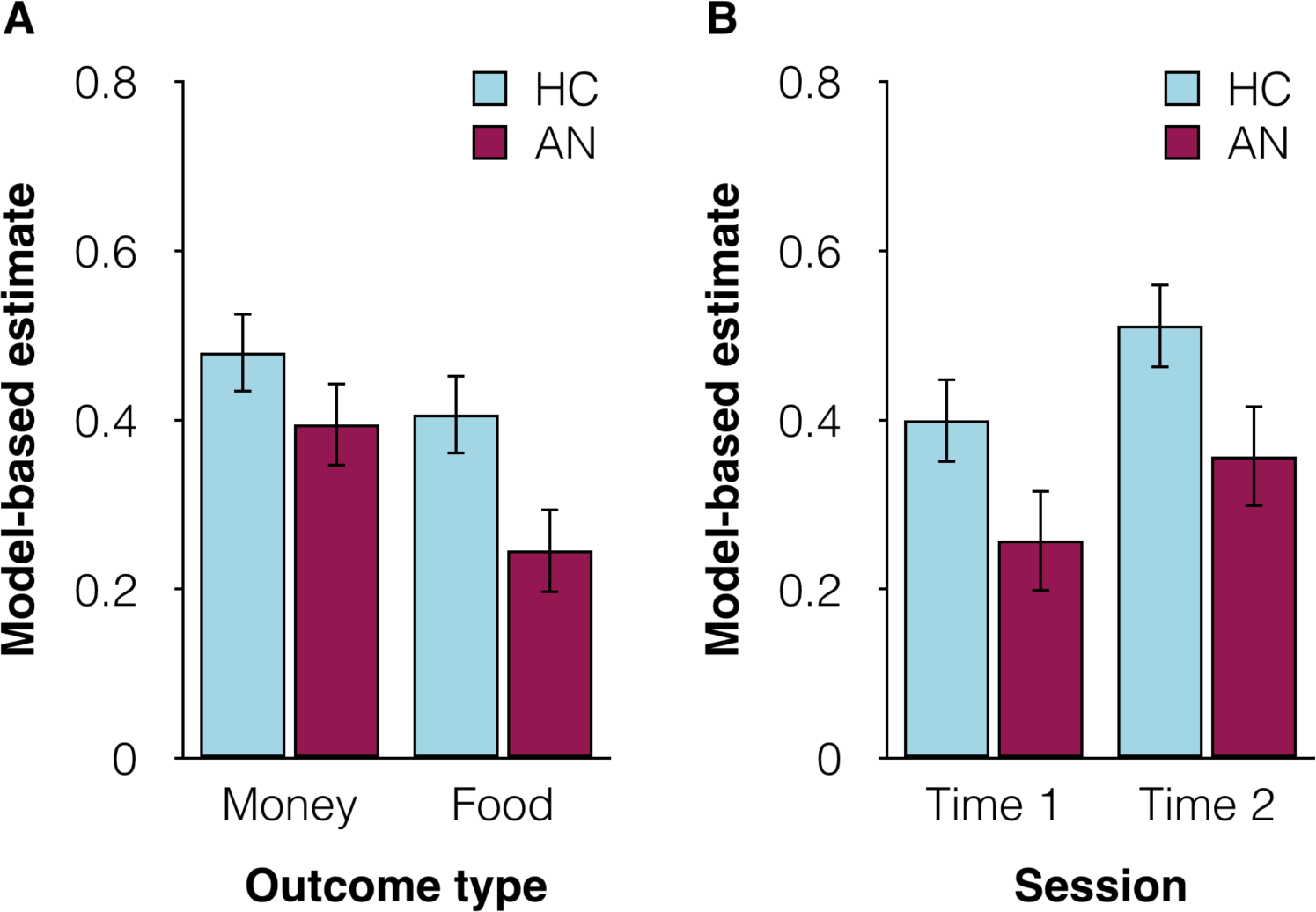
**A**) Model-based contributions to learning for the monetary and food tasks across session (Time 1 and Time 2). **B**) Model-based contributions to learning at Time 1 and Time 2 across task type (monetary and food task).

The contribution of model-based vs. model-free learning may be hypothesized to differ based on the state of illness. At Time 1, patients were generally undernourished (with potential sequelae for neurocognitive functioning) and had higher psychological symptom burden (e.g., depression and anxiety, see **Table 1**). Therefore, we tested participants before (Time 1) and after (Time 2) weight restoration treatment. Although there appeared to be a tendency toward more model-based behavior at Time 2 in both groups, this main effect was not significant (p = 0.15) and there was no interaction between group and Session (Time 1 vs. Time 2) on model-based learning (Est: 0.02, SE = 0.08, z = 0.27, p = 0.79, **Fig. 3B)**.

### Post-Test Assessment of Task Transition Structure

HC and AN did not differ significantly in their recall of the transition structure for the monetary or food outcomes task at Time 1 (Money: χ^2^(1, *N* = 87) = 0.274, *p* = 0.60; Food: χ^2^(1, *N* = 84) = 0.925, *p* = 0.34) or Time 2 (Money: χ^2^(1, *N* = 52) = 0.006, *p* = 0.94; Food: χ^2^(1, *N* = 52) = 0.650, *p* = 0.42). HC and AN also did not differ significantly on the average discrepancy between their estimates of the transition probabilities and the actual probabilities at Time 1 (Money: t(82) = -0.91, p = 0.37; Food: t(82) = -0.59, p = 0.56) or Time 2 (Money: t(51) = -1.73, p = 0.09; Food: t(51) = -1.34, p = 0.18).

### Correlations with clinical variables

Model-based behavior was not significantly associated with BMI (over and above the group difference) or duration of illness in the patient group (BMI: Est: 0.003, SE = 0.03, z = 0.09, p = 0.93; Duration: Est: 0.05, SE = 0.07, z = 0.80, p = 0.42). In AN at Time 1, higher scores on the YBC-EDS were associated with less model-based learning for food outcomes relative to monetary outcomes (Est: 0.09, SE = 0.05, z = 1.95, p = 0.051), a trend mainly driven by (and significant for) the Preoccupations subscale (Est: 0.13, SE = 0.04, z = 3.07, p = 0.002) rather than the Rituals subscale (Est: 0.04, SE = 0.05, z = 0.77, p = 0.44). (Data were not collected from HC, for whom scores would be expected to be zero, or from AN after weight restoration.)

### AN subtypes

Individuals with AN can be classified as either restricting (AN-R) or binge eating/purging (AN-BP) subtype and the current sample was split approximately equally between subtypes (19 AN-R, 22 AN-BP). Exploratory analyses suggested that the impairment in model-based learning was largely driven by individuals with AN meeting criteria for binge-eating/purging, rather than restricting subtype (see **supplementary material** and **supplementary Fig. S1**).

## Discussion

This study addressed three questions about the intersection of computational neuroscience and psychiatry by examining goal-directed and habitual learning mechanisms in AN. We tested whether AN, which appears at once excessively goal-directed *and* habitual, is characterized by enhanced or diminished model-based behavior. We further examined model-based versus model-free learning in a food-specific context as well as a monetary context, to test the domain-specificity of any differences, and before and after weight restoration, to test whether impairments were present only during acute illness or were instead a more stable characteristic of patients with AN. Individuals with AN showed less model-based learning than HC, and groups did not differ in model-free learning. This group difference was present when playing for both food and monetary outcomes and persisted after successful weight-restoration treatment.

### How does the extreme, yet inflexible, “self-control” that characterizes AN relate to model-based learning?

AN poses a fascinating conceptual challenge for the classic psychological dichotomy between goal-directed, controlled behavior and inflexible, habitual responses. Selection of a low-fat, low calorie diet with limited variety is a stereotyped feature of illness in AN (Mayer, Schebendach, Bodell, Shingleton, & Walsh, 2012; Schebendach et al., 2008; Sysko, Walsh, Schebendach, & Wilson, 2005). Yet nearly half of patients also engage in eating that is accompanied by a sense of loss of control (binge eating). The coexistence of these behaviors, as well as other indicators that behavior is actually quite inflexible in AN, makes this illness particularly intriguing as an opportunity to better understand the relative contributions of goal-directed and habitual behavior. Maladaptive behaviors in AN are commonly understood to reflect heightened self-control, which might be expected to relate to enhanced dominance of goal-directed (or model-based) behavior. Yet these same behaviors are also rigid and difficult to change, which might suggest the opposite result: relatively weakened model-based control. This study allowed us to test existing hypotheses that impairments in goal-directed, or model-based, behavior are a transdiagnostic mechanism underlying compulsive psychopathology (Gillan et al., 2016; Voon et al., 2015).

Our results suggest that, compared to HC, AN rely significantly less on the model-based approach, similar to individuals with other disorders involving compulsion such as OCD or drug abuse (Gillan et al., 2016; Voon et al., 2015). The pathology of AN involves complex, multi-step phenomena to avoid food intake. The findings in this study suggest that these computational mechanisms may be relevant even to producing behaviors, like pathological food avoidance, that extend beyond the traditional notion from animal behavioral psychology of habits as simple stimulus-response motor programs. Indeed, a recent study in a large general population sample found that model-based behavior in the Two-Step task, used herein, correlated with a set of psychiatric symptoms that included not just simple compulsive actions (such as repetitive checking or morning drinking) that extend easily from a basic notion of habits but also broader, more cognitive symptoms such as intrusive thoughts (Gillan et al., 2016). In contrast, this study found no significant difference between AN and HC in the model-free approach in the task, and groups also differed significantly in a composite index of relative reliance on the two systems, supporting the specificity of the effect. That said, as with previous studies using this task (Gillan et al., 2016), we focused our hypotheses and analyses on the modulation of model-based behavior, as this has proven to be the measure from this task that has most reliably tracked manipulations or individual differences likely to be relevant to the goal-habit balance (Otto, Raio, et al., 2013; Otto, Skatova, Madlon-Kay, & Daw, 2015).

### Does specific psychopathology reflect a domain-general failure of model-based learning?

Our data indicate that the deficit in model-based learning in AN was not significantly remediated by using food outcomes. Rather, it was consistent across both domain-general (money) and illness-specific (food) outcomes, with no significant task differences in the group effect. This finding cuts against one potential explanation for the current results on the money task and other similar results showing reduced model-based learning for money in psychiatric populations (Voon et al., 2015): that generic monetary outcomes are less motivating in psychiatric populations than they are for HC participants. Interestingly, it has recently been shown that incentives *can* reliably boost model-based performance–even in the more severe end of the spectrum for a range of psychopathology constructs (Patzelt, Kool, Millner, & Gershman, 2019). By using an incentive-compatible food outcome (success on the Two-Step task led to actual snack choices after the task), the present study included a condition in which motivation would be expected to be more salient for individuals with AN than for HC. Yet, even in the food context, patients with AN showed less model-based behavior than HC. Of course, the failure to detect a Group-by-Task interaction is a negative result, and as with any such result, the possibility remains that there does exist some interaction smaller than our study was powered to detect. However, this is mitigated by a positive result: that AN were significantly impaired at model-based learning, relative to HC, even in the food task considered alone (Est=0.17, SE=0.067, z=2.6, p=0.01).

Exploratory analyses of the relationship between symptom severity and task performance in AN did suggest one result that was selective for the food task: higher scores on the YBC-EDS, especially the Preoccupation subscale, were associated with less model-based learning for food outcomes relative to monetary outcomes. Such a graded deficit seems plausible, and its specificity to food intriguing, but it is of note that the direction of the effect (worse learning with food outcomes) is opposite that predicted by a motivational account. Also, given the exploratory nature of the analysis and the smaller number of data points underlying it, there is a danger of false positives and replication will be required to confirm this finding.

### Are model-based deficits secondary to psychopathology, or potentially primary?

The idea that compulsive psychopathology might relate to impaired model-based learning is appealing in part because it hints at a causal mechanism by which pathological habits might emerge. Of course we are not in a position to address causality, nor even whether the cognitive deficits precede the symptoms. However, we were able to address whether the cognitive differences between AN and HC persisted after acute weight restoration. That they did indicates they were not due to starvation alone. Prior research has shown that while psychological measures improve with weight restoration, eating behavior—namely, the pursuit of low-fat low-calorie diets—does not improve substantially with weight restoration (Mayer et al., 2012; Sysko et al., 2005). Additionally, in a previous study we found that deficits in feedback-based learning remained after weight restoration (Foerde & Steinglass, 2017). The effect on model-based learning here shows a similar persistence, consistent with model-based deficits as a relatively stable characteristic in AN. This result also parallels a recent report, using the same task, that cognitive behavioral therapy for OCD also does not improve goal-directed deficits in that population (Wheaton et al., 2018). In the current dataset, the absence of an effect of weight restoration is a negative result and subject to similar interpretational caveats as the money vs. food contrast. However, the confidence interval on the effect suggests that any undetected effect of weight restoration on model-based behavior is likely to be quite small relative to the overall deficit.

## Conclusion

Previous work has shown deficits in responding to reward (DeGuzman, Shott, Yang, Riederer, & Frank, 2017; Frank, Shott, Hagman, & Mittal, 2013; O’Hara, Campbell, & Schmidt, 2015) and learning from feedback in AN (Foerde & Steinglass, 2017; Lawrence et al., 2003; Shott et al., 2012). The present results suggest that a more fine-grained parsing of this deficit is necessary. Basic habitual learning mechanisms may be intact, whereas flexible responses to changing contingencies and the ability to integrate a model of the environment into choices are impaired. Individuals with AN understood the structure of the task as well as did HC, as both groups were able to report verbally on the rules of transition in the task. Yet, individuals with AN were less able to successfully use this information in their decision-making during the task. These findings are consistent with the habit-centered model of AN (Walsh, 2013), which argues that, once established, patterns of maladaptive eating in AN are no longer goal-directed. Relatedly, maladaptive food choice is mediated in AN by dorsal striatum (Foerde et al., 2015).

Nonetheless, a paradox remains in that individuals with AN are characterized by less model-based behavior but also more patient delay discounting in inter-temporal choices (Decker et al., 2015; Steinglass et al., 2012; Steinglass et al., 2017). Generally, decreased model-based behavior would be expected to be associated with increased delay discounting: both are broadly associated with impulsivity, both are related to measures of executive functioning (Otto et al., 2015; Shamosh et al., 2008), and both are enhanced by increasing dopamine levels in patients with Parkinson’s disease (Foerde et al., 2016; Sharp, Foerde, Daw, & Shohamy, 2016). Also, a recent study found that they were related across individuals (Hunter, Bornstein, & Hartley, 2018). One potential resolution of the paradox could lie with further consideration of subtypes of AN. The present study found that the deficit in model-based learning was primarily present in the AN-BP rather than the AN-R subtype (a pattern also found in a previous study, (Foerde & Steinglass, 2017)). In contrast, decreased delay discounting has primarily been seen in the AN-R rather than the AN-BP subtype (Decker et al., 2015; Steinglass et al., 2012). Further explication of these patterns will rely on larger samples suited to parse the contributions of subtypes and computational neuroscience and psychiatry approaches may be fruitful in understanding the distinct manifestations of psychopathology within complex illnesses such as AN.

The divergence between goal-directed and habitual behavior in AN could have implications for how illness develops and how it becomes resistant to change. It is particularly important for an illness that often develops in adolescence, a time during which model-free learning reigns and model-based behavior only begins to emerge (Decker et al., 2016). The slower emergence of model-based behavior across development suggests one mechanism that may nudge dieting behavior down the habit (model-free) path, and may be one contributing factor to how this eating behavior becomes so entrenched. More generally, these results also speak to the promise of the goal-directed vs habitual dichotomy as a trans-diagnostic mechanism, associated with compulsive symptomatology across disorders: the decision-making deficit here was domain-general, not food-specific, and apparently not secondary to starvation. Finally, they speak as well to the architecture of decision making in the healthy brain: they suggest that the sorts of behaviors emitted in the absence of control go well beyond simple stimulus-response habits, speaking to the inadequacy of current psychological and computational conceptions of automatic behavior.

In summary, we found that model-based, and not model-free, behavior was impaired among individuals with AN relative to HC. The deficit was not remediated by using real food as the outcomes of choices to enhance motivational salience nor by weight restoration and associated improvement in psychological symptoms.

## Materials and Methods

### Participants

Participants were 41 women with AN and 53 healthy comparison women (HC). Individuals were eligible if they were between the ages of 16 and 46 years (**Table 1**), with an estimated IQ > 80 (measured by Wechsler Abbreviated Scale of Intelligence, WASI-II, (Wechsler, 1999)). Eligible patients met DSM-5 (American Psychiatric Association, 2013) criteria for AN—restricting (AN-R, n=19) or binge eating/purging (AN-BP, n=22) subtype—and were receiving inpatient treatment at the New York State Psychiatric Institute (NYSPI) specialized Eating Disorders Unit. Patients with AN were not eligible if they had a history of a psychotic disorder, were at imminent risk of suicide, or met criteria for substance use disorder. Anxiety and depressive disorders were not exclusionary, as these commonly co-occur with AN (Hudson, Hiripi, Pope, & Kessler, 2007). Three individuals with AN were taking antidepressant medications (SSRIs). HC were recruited through the community and were compensated $50 for their time. HC were group-matched for age and ethnicity and were included if they had no current or past psychiatric illness, including any history of an eating disorder, and had a body mass index (BMI) in the normal range (18–25 kg/m^2^). The HC group included 19 individuals who endorsed a history of dieting behavior. This study was approved by the NYSPI Institutional Review Board; after complete description of the study to the participants, adult participants provided written informed consent and adolescents provided written assent with parental consent.

### Procedure

Psychiatric diagnoses were established using the Semi Structured Interview for DSM-IV (SCID; (First, Spitzer, Gibbon, & Williams, 2002) and the Eating Disorders Assessment for DSM5 (EDA-5, (Sysko et al., 2012)). Height and weight were obtained on a beam balance scale (Detecto, Webb, MO). Estimated IQ was assessed with the WASI-II (Wechsler, 1999). Severity of eating disorder psychopathology was measured by the Eating Disorder Examination Questionnaire (EDE-Q; (Fairburn, 2008; Fairburn & Beglin, 1994)), a 36-item self-report assessment of eating disorder symptoms which has established community norms for adolescents and adults. The four symptom subscales (Restraint, Eating Concern, Shape Concern, and Weight Concern) are scored on a scale of 0-6 and averaged to obtain a “global” score. In addition, participants with AN completed the Yale-Brown-Cornell Eating Disorder Scale (YBC-EDS; (Mazure, Halmi, Sunday, Romano, & Einhorn, 1994)), an interview measure of eating disorder symptoms with separate subscales related to Preoccupations and Rituals. Each subscale is scored on a scale from 0-16 and the subscales are summed to obtain the total score (ranging from 0-32). Higher scores indicate greater symptom severity on each measure.

Participants were assessed at two points: Individuals with AN were tested within 1 week of hospital admission (Time 1) and again after inpatient weight restoration treatment to at least 90% ideal body weight (Metropolitan Life Insurance, 1959), corresponding to a body mass index (BMI) of approximately 19.5-20.0 kg/m^2^ (Time 2). HC individuals were tested twice at an interval group matched to AN (M_HC_ = 51 +/- 23 days, M_AN_ = 59 +/- 22 days, t(45) = 1.17, p = 0.25). Of 41 AN participants, 18 completed both time points, 16 completed only Time 1, and seven completed only Time 2; of 53 HC participants, 29 completed both time points and 24 completed only Time 1. For one AN participant, data from the food outcome task at Time 2 was lost due to computer malfunction.

### Two-Step Decision Task

Each participant completed two versions of the Two-Step Decision Task (Daw et al., 2011; Decker et al., 2016; Sharp et al., 2016)—one that involved playing for money and one that involved playing for a food snack to be consumed after the task (order counterbalanced across participants). The tasks were structurally identical but included distinct cover stories and task environments that matched the outcomes used (see **Fig. 1AB**). For both tasks each trial proceeded in two stages (**Fig. 1C**). In the first stage (Stage 1), participants chose between two spaceships (or cafés), revealing a second-stage (Stage 2) choice between two aliens (or food trays). Each second-stage alien (or food tray) had a slowly changing chance of delivering space treasure (or a food token) vs. nothing, necessitating continuous learning by trial and error. The four Stage 2 options were determined by independently drifting Gaussian random walks with a standard deviation = 0.025 and bounded by 0.25 and 0.75 probability of reward, such that the reward probability associated with each Stage 2 option changed slowly from trial to trial (**Fig. 1D**). The response window at each stage was three seconds. Participants completed 200 trials of each task.

A key design feature of the task was the probabilistic association between first- and second-stage choices: Choosing the blue spaceship (or blue café) lead to the purple planet (or green kitchen) 70% of the time—a *common* transition, and the red planet (yellow kitchen) 30% of the time—a *rare* transition. The contingencies were reversed for the green spaceship (pink café). The transition structure between stages allows dissociation of model-free vs. model-based learning strategies: Model-free learning is ignorant of transition structure and favors repeating a first-step choice that ultimately results in reward, even if it does so via a low probability transition. By contrast, model-based learning is sensitive to the transition contingencies and uses them to infer the first-stage choice most likely to lead to the preferred second-stage environment.

In the monetary version of the task, each piece of space treasure collected was worth $0.10 and the total earned was paid out in cash at the end of the experiment. In the food version of the task, the accrued tokens bought access to a selection of foods from which to choose a snack item. Prior to completing the task, participants were given a “menu” (**supplementary Fig. S2**) with pictures of 15 food items and were asked to rank them in order of preference, but were not told that the ratings were related to later snack options. An increasing number of earned food tokens granted access to an increasing number of preferred food items (e.g., 92 tokens provided access to items ranked 9-15, 114 tokens or more provided access to all items, see **supplementary Table S1**).

A subset of participants (HC: n=9, AN: n=18) completed a different version of the food decision task (the monetary task was the same for all participants). In this version, rather than receiving a food token vs. nothing as the second-stage outcome, participants saw a picture of a preferred food item vs. a non-preferred food item. The specific food items were individually determined per participant: prior to the decision task participants completed a rating task in which 9 food items were rated on a scale from 1-9 (1= highly preferred). Participants were not instructed that their ratings would relate to their later snack options. The food image participants received on most trials determined which snack they would receive after the task. In this task, participants had 2 seconds to make choices in both stages. Results were qualitatively similar for both food tasks, and they were therefore combined in all analysis below.

After completing the Two-step tasks, participants’ knowledge of the transition structure between Stage 1 and Stage 2 (e.g., *If you picked the blue spaceship, which planet would you most likely land on?)* and their estimates of the transition probabilities were assessed (e.g., *If you picked the blue spaceship, how likely would you be to see the purple planet?)*. Responses were not collected from 3 HC for the monetary task at Time 1 and from 1 AN on both the monetary and food tasks at Time 2.

## Data analysis

Demographic and clinical variables were compared using independent t-tests. Measures of psychopathology at Time 1 and Time 2 were compared within AN using paired t-tests.

### Assessment of Model-based and Model-free learning (Computational Model)

In order to capture the influence of incremental learning across many trials, we fit subjects’ choices using a reinforcement learning model in which choices are modeled as arising due to the weighted combination of model-free and model-based reinforcement learning. The model is equivalent to the one used by Gillan et al. (Gillan et al., 2016) and Otto et al. (Otto, Raio, et al., 2013), except that following Sharp et al. (Sharp et al., 2016) we have eliminated one free parameter (by setting *β*^*MF*0^ in the GIllan model to 0, or equivalently *λ* in the Otto model to 1). It reflects a simplification and a change of variables over the model from Daw et al. (Daw et al., 2011).

On each trial, *t*, participants make a Stage 1 choice *c*_1,*t*_, leading to a transition to a Stage 2 state *s*_*t*_ where another choice, *c*_2,*t*_ is made, followed by reward *r*_*t*_. At Stage 2, it is assumed that participants learn a value function over states and choices, *Q*^*stage*2^(*s, c*), whose value for the chosen action is updated based on the reward received at each trial according to a delta rule, 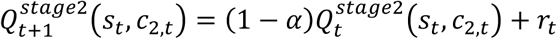. Here, *α* is a free learning rate parameter. (In this, and analogous update equations, a factor of *α* is omitted from the last term of the update, equivalent to rescaling the rewards and *Q*s by 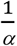 and the corresponding weighting parameters *β* by *α* (Otto, Raio, et al., 2013)). The probability of a particular Stage 2 choice is modeled as governed by these values according to a logistic softmax, with free inverse temperature parameter 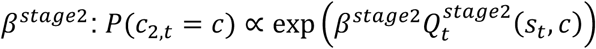, normalized over both options *c*.

Stage 1 choices are modeled as determined by the weighted combination of both model-free and model-based value predictions about the ultimate, Stage 2 value of each Stage 1 choice. *Model-based* values *Q*^*ME*^ are given by the learned values of the corresponding Stage 2 state, maximized over the two actions: 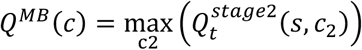, where *s* is the Stage 2 state predominantly produced by Stage 1 choice *c. Model-free* values are learned by TD(1), where 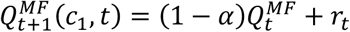. The Stage 1 choice probabilities are given by a logistic softmax, with a contribution from each value estimate weighted by its own free inverse temperature parameter: 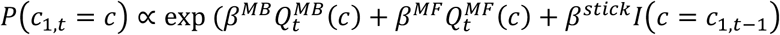. Here, *I*(*c* = *c*_1,*t*−1)_ is a binary indicator of whether a choice repeats the previous trial’s choice, so the weight *β*^*stick*^ measures the general tendency to perseverate or switch regardless of feedback.

At the end of each trial, all value estimates *Q* for unchosen actions and unvisited states are decayed by multiplying by (1 − *α*).

The model has a total of five free parameters: four weights *β* (*β*^*stick*2^, *β*^*MB*^, *β*^*MF*^, *β*^*stick*^ and a learning rate *α*. Our main measures of interest are *β*^*MB*^ and *β*^*MF*^, measuring the contribution of model-based and model-free learning.

The free parameters of the model were estimated by maximizing the likelihood of each participant’s sequence of choices, using a distinct set of parameters for each game (i.e., each combination of participant, session, and task type: up to four games per participant, two task types at two timepoints). These were estimated jointly with group-level distributions over the entire population using an Expectation Maximization procedure (Huys et al., 2011) implemented in the Julia language (Bezanson, Karpinski, Shah, & Edelman, 2012). The per-game model-based and model-free weightings *β*^*MB*^ and *β*^*MF*^, indicating the strength of each type of learning, were extracted for further analysis. (We also computed a standard measure of their relative fractional reliance, 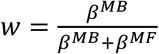; Daw et al, 2011.) These estimates were used as dependent variables in a series of regression analyses with group as the main explanatory variable of interest. All analyses controlled for task type, session, IQ, and age as additional independent variables. In followup analyses, we included interactions of group by session or task, and measures of disease severity (BMI, duration illness, and YBC-EDS); all covariates were z-scored. The regressions were conducted using mixed-effects logistic regression, and estimated using Julia’s MixedModels package. All within-subject parameters (e.g. task and session) were taken as random effects per participant, so as the to capture the repeated-measure structure of the data (e.g., tasks repeated at two timepoints) and also the imbalanced data (e.g., not all subjects completed both timepoints).

## Data Availability

Data will be available upon reasonable request once the manuscript has been published.

## Supplementary Information for

### Model-based performance in AN-R vs. AN-BP subtypes

We further assessed model-based performance separately for ANs meeting criteria for restricting (AN-R) vs. binge-eating/purging (AN-BP) subtype. As seen in Figure S1, HC were most model-based, and AN-BP least model-based, with AN-R falling in between. The HC and AN-BP groups differed significantly from each other (Est: 0.21, SE = 0.06, z = 3.36, p = 0.0008, whereas AN-R did not differ significantly from AN-BP (Est: 0.12, SE = 0.09, z = 1.33, p = 0.18) or HC (Est: -0.09, SE = 0.08, z = -1.05, p = 0.29).

**Figure S1:**
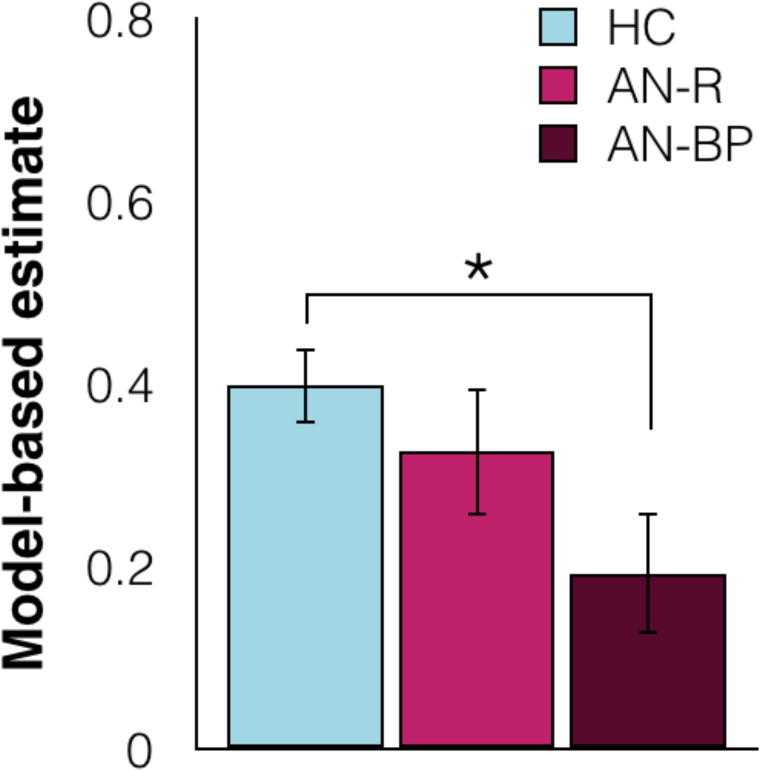
Overall model-free contributions to learning for healthy controls (HC), AN with restricting subtype (AN-R) and AN with binge-purge subtype (AN-BP) across task type (monetary and food) and session (Time 1 and Time 2).

**Figure S2:**
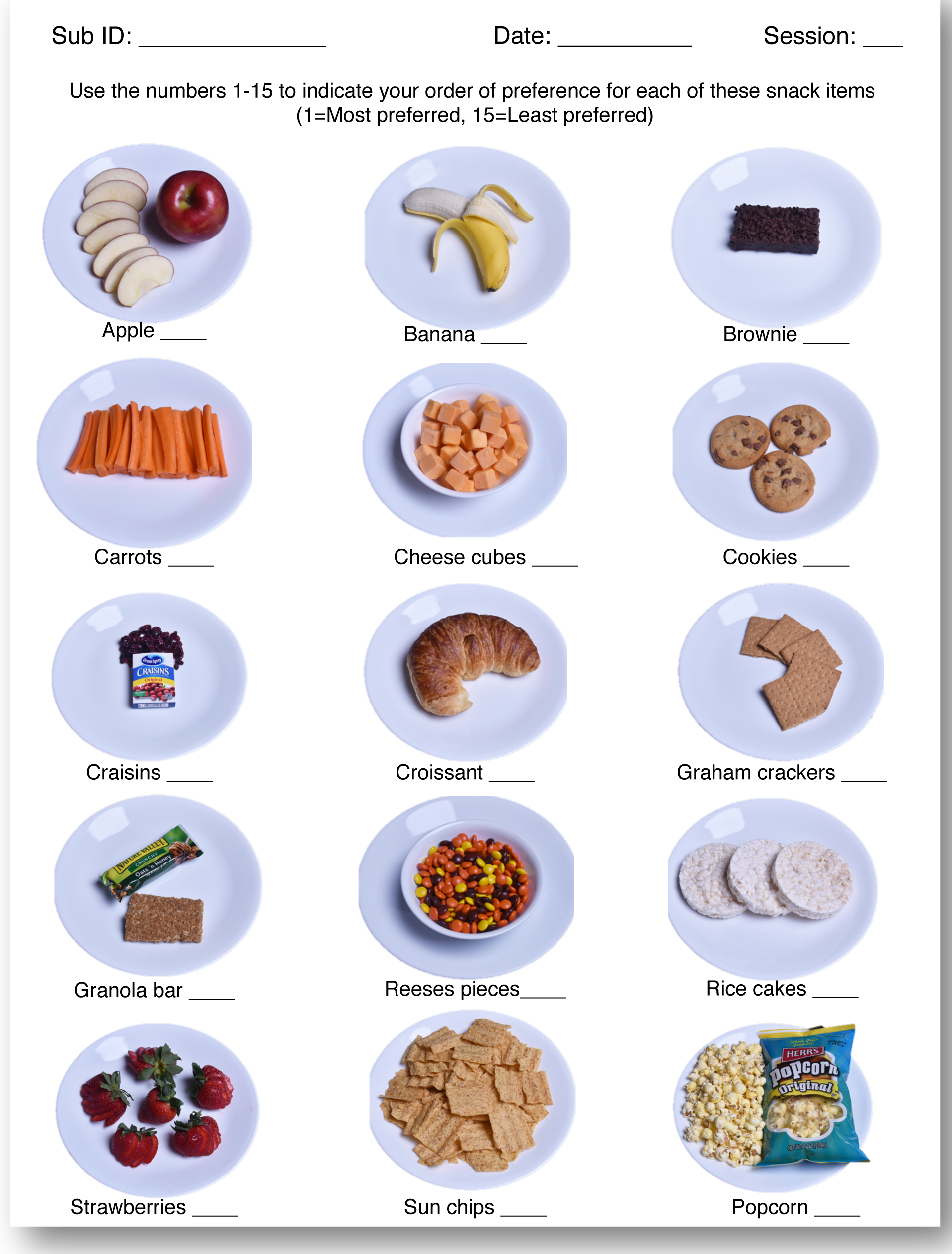
Food task menu used to determine outcomes for the food version of the 2-step task. Participants numbered items in order of preference. Performance on the task determined access to food items, with better performance granting access to increasingly preferred items (see **Table S1**).

**Table S1.** Conversion of performance into snack options

| Performance |  | Access to snack<br>item rank |
| --- | --- | --- |
| (proportion correct) | (tokens earned) |  |
| $\geq 0.57$ | $\geq 114$ | 1<sup>st</sup> |
| 0.55 – 0.569 | 110 | 2<sup>nd</sup> |
| 0.54 – 0.549 | 108 | 3<sup>rd</sup> |
| 0.53 – 0.539 | 106 | 4<sup>th</sup> |
| 0.52 – 0.529 | 104 | 5<sup>th</sup> |
| 0.51 – 0.519 | 102 | 6<sup>th</sup> |
| 0.50 – 0.509 | 100 | 7<sup>th</sup> |
| 0.49 – 0.499 | 98 | 8<sup>th</sup> |
| 0.46 – 0.489 | 92 | 9<sup>th</sup> |
| $< 0.46$ | $< 92$ | 10<sup>th</sup> |

